# Care Delivery Outcomes of an Early Pregnancy Access Center

**DOI:** 10.64898/2026.05.18.26353517

**Authors:** Sameera Mokkarala, Alice Abernathy, Nathanael Koelper, Arden McAllister, Sarita Sonalkar, Courtney Schreiber

## Abstract

**Objectives:** To evaluate if direct access to a Pregnancy Early Access Center (PEACE) improves the timeliness and efficiency of pregnancy loss care.

**Methods:** We conducted a retrospective cohort study of patients diagnosed with EPL from January 2017 to December 2022 within a single healthcare system. We included EPL patients treated with procedural or medication management who had been assessed for a related early pregnancy complaint in the thirty days prior. The exposure was direct utilization of PEACE (yes/no) between first EPL symptom visit and EPL management. The primary outcome was “care latency” defined as days from initial presentation for concerning early pregnancy symptoms to initiation of active management. Secondary outcomes included “care continuity,” the number of care teams encountered, “care efficiency,” the number of patient encounters, and the type of EPL management received.

**Results:** The evaluable cohort included 2151 individuals, with 36.5% patients of Black race and 30.3% publicly insured. A total of 885 (41.1%) received any EPL care at PEACE and 246 (11.4%) initiated their care at PEACE. Patients initiating care through PEACE experienced a 5-day reduction in care latency compared to patients who did not access PEACE. Adjusting for age, race, and insurance type, patients whose index EPL visit was with PEACE initiated their treatment twice as quickly as those who never saw PEACE (aHR 2.36 [95% CI, 2.05-2.71]). Care efficiency (median 2 [1–3] encounters) and care continuity (median 4.5 [4–7] care teams) were also improved by an index visit with PEACE when compared with controls (3 [2–4] and 6 [4–8] p<0.01), respectively).

**Conclusions:** The Pregnancy Early Access Center (PEACE) model is associated with reduced care latency and improved efficiency and continuity when compared with routine care. PEACE reduces barriers to timely, patient-centered early pregnancy care.

## Introduction

Early pregnancy loss (EPL) affects approximately 20% of pregnancies[1]. EPL management can occur in various settings. However, for reasons including lack of an established provider early in pregnancy[2] and current obstetrical reimbursement models, EPL-related visits account for over 900,000 emergency department (ED) visits annually[3] in the United States (US). In the post-Dobbs environment, fragmentation of reproductive healthcare access may further exacerbate delays in evaluation and management of early pregnancy complications.

ED utilization is more prevalent among vulnerable populations, partly due to ambulatory access barriers[4]. Patients with EPL seen in the ED are more likely to report negative care experiences compared to those seen in outpatient settings[4–7]. While efforts are underway to improve the quality of ED-based EPL care[8], alternative models are evolving.

Early Pregnancy Units, first implemented in the United Kingdom in 1991, are ambulatory clinics that specialize in diagnosing and managing early pregnancy complications, including EPL and ectopic pregnancies. This model improves care quality and satisfaction for those with concerning early pregnancy symptoms[9–11]. Expanding access to early pregnancy assessment clinics in the US could reduce ED visits for EPL and provide more patient-centered and comprehensive care[12,13].

Our model, the Pregnancy Early Access Center (PEACE), was established and refined between 2014 and 2017 at Penn Medicine, Philadelphia, PA. In this study, we assessed whether receiving care with PEACE improved the patient experience by reducing the duration of time between seeking evaluation and active management treatment, the total number of clinical encounters required, and the number of care teams encountered. We additionally examined the demographic characteristics associated with accessing PEACE care.

## Methods

### Study Design and Cohort Definition

We conducted a retrospective cohort study of pregnant patients who experienced EPL that was actively managed (i.e., received uterine aspiration or medication management) within our urban healthcare system between January 2017 and December 2022, deemed exempt by the University of Pennsylvania Institutional Review Board. We used *International Classification of Diseases, Ninth or Tenth Revision, Clinical Modification* (ICD-9/10-CM) diagnosis codes (Appendix 1) to identify patients with an EPL diagnosis made during the study period. Of patients with an EPL diagnosis, we included in our cohort patients who 1) had a visit within the health system for evaluation of a concerning pregnancy symptom (bleeding or cramping) in the thirty days prior to EPL diagnosis entry, and 2) a billing or procedure code for EPL management with medication or uterine aspiration entered within the sixty days following EPL diagnosis code entry (Fig 1). Because expectant management cannot be reliably identified through administrative coding, the study focused on patients receiving active management.

**Figure 1.**
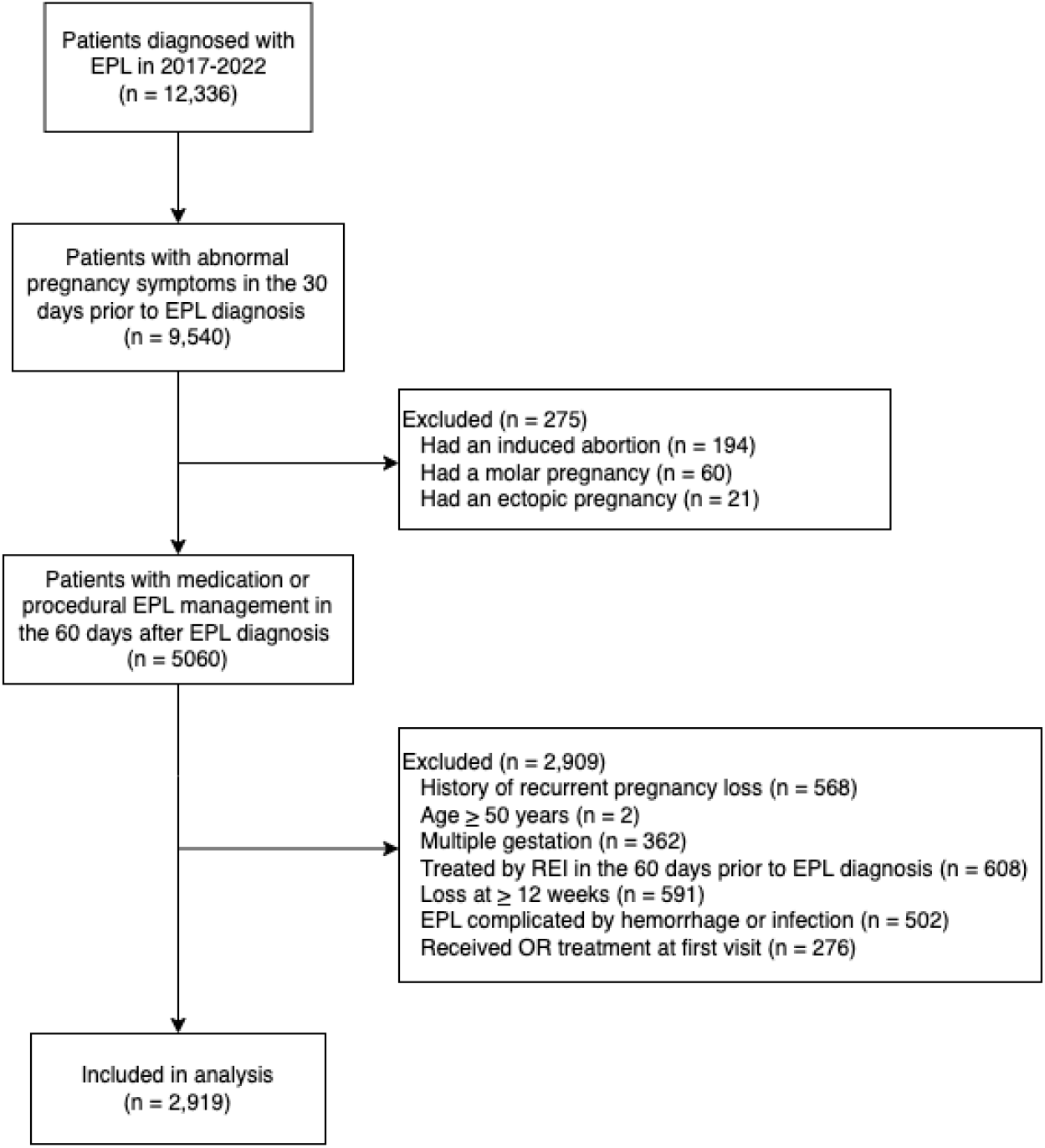
Flow diagram of study cohort identification. EPL, early pregnancy loss; REI, reproductive endocrinology and infertility; OR, operating room.

**Figure 2.**
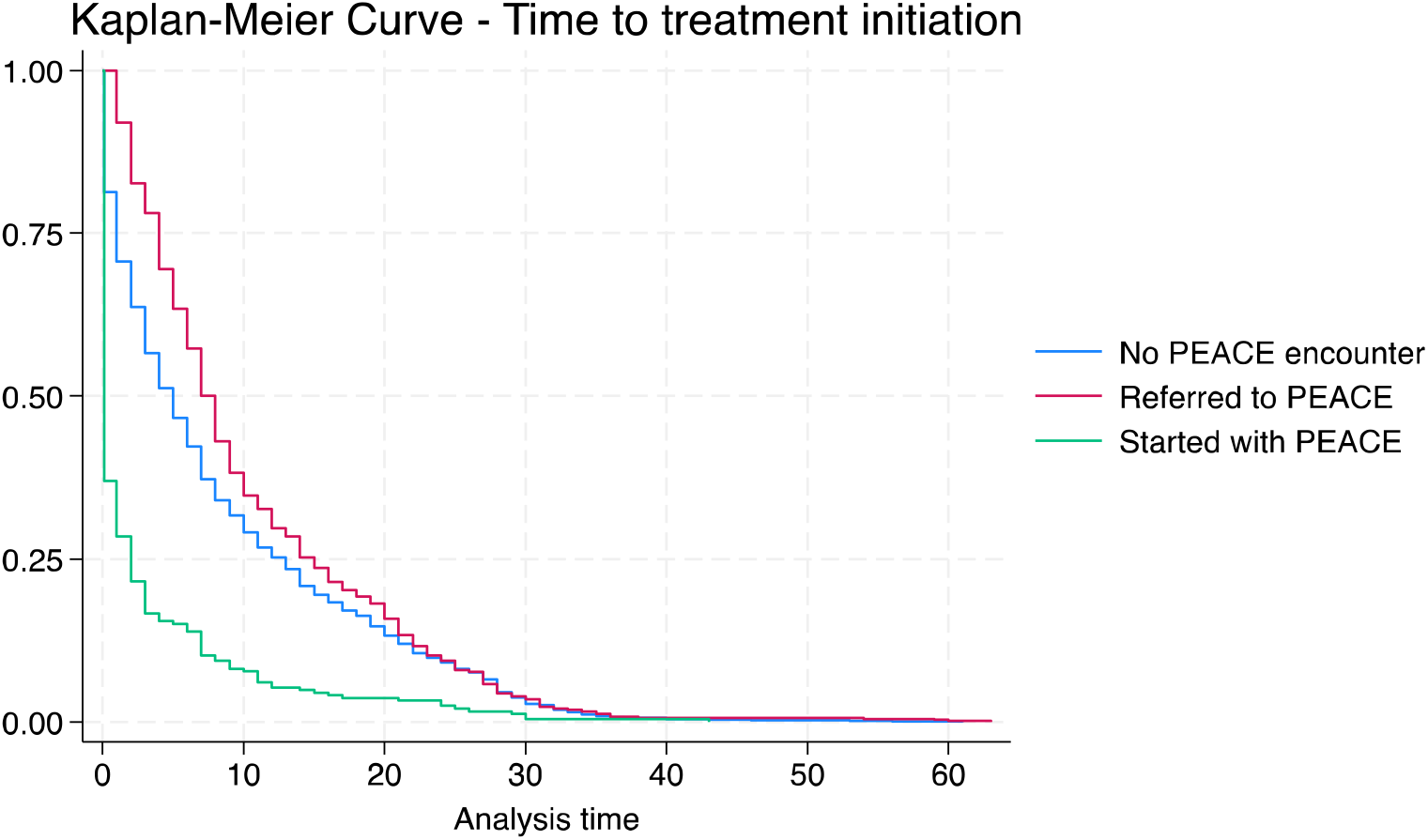
Time to treatment initiation for patients by PEACE (Pregnancy Early Access Center) utilization category

Within our healthcare system, patients can receive EPL care in one of four ways: 1) through the ED without ambulatory care, 2) with a general gynecology provider, 3) with PEACE directly (i.e., through self-referral), or 4) with PEACE via referral from another provider. In this study, we aimed to compare the outcomes between ambulatory sites, and therefore did not include those who received their care directly through the ED. Across the three ambulatory options for management, care is available during standard business hours (Monday through Friday, 8:00AM to 5:00PM). PEACE offers services including early pregnancy evaluation and complication management, provided by a team of physicians and nurse practitioners. Bedside ultrasound, medication administration and prescription, and uterine aspiration under local anesthesia are available on-site. Patients who prefer or require procedural management under sedation and are deemed to be clinically stable can be scheduled for care in an ambulatory surgical facility.

Among those patients actively managed, exclusion criteria included patients with diagnosis codes for ectopic and molar pregnancies, induced abortions, and pregnancy losses occurring at greater than twelve weeks and zero days of gestation. We additionally excluded patients who conceived using assisted reproductive technology (defined as subjects who had an encounter with reproductive endocrinology and infertility in the sixty days prior to their EPL diagnosis), as these patients generally have facilitated care access until hand-off to routine prenatal care. Additionally, we excluded patients with diagnoses indicating clinical instability including “hemorrhage”, “infection”, “endometritis”, or “sepsis” during the ninety day study period, as these patients would have been ineligible for non-urgent management and therefore would not have been referred for ambulatory care at PEACE or elsewhere. A random sample of the cohort was reviewed to ensure actively managed, stable EPL cases were accurately captured.

For patients meeting inclusion criteria, we obtained demographic information; the contact date and department specialty for each in-person clinical utilization with an associated EPL-related diagnosis code that occurred during the patient’s inclusion period; and the date of treatment initiation. EPL management types were categorized into three groups: (1) medication management, defined by prescription of mifepristone 200 mg and misoprostol 800 mcg, (2) in-office uterine aspiration, defined by the ordering of both oral doxycycline 200 mg and the appropriate uterine aspiration procedure code, and (3) operating room (OR) uterine aspiration under sedation, defined by the ordering of intravenous doxycycline 200 mg, the appropriate uterine aspiration procedure code, and a co-occurring clinical encounter with anesthesia (Supplemental Table 1). Inclusion of the route of doxycycline administration allowed for confirmation of the procedure’s completion, as well as differentiation between the office setting (in which oral doxycycline is given) and OR setting (in which intravenous doxycycline is given) to reduce misclassification bias. For patients who received multiple treatments during the study period, the first management type received was considered the treatment employed.

### Exposures

We defined the exposure as any in-person utilization of PEACE occurring during a patient’s inclusion period. We stratified PEACE patients into two groups: 1) those who encountered PEACE as the index point of care and 2) those who came to PEACE at any point during their care by way of referral. A third group, those who did not encounter PEACE at all during their inclusion period, served as “routine care” controls.

### Outcomes

Our primary outcome was “care latency,” defined as the number of days between initial presentation for evaluation of a concerning pregnancy symptom and their visit for EPL management initiation. Secondary outcomes included “care efficiency,” defined as the total number of clinical encounters the patient has during their inclusion period, and “care continuity,” defined as the number of different specialty teams or unique departments the patient encountered during their inclusion period (for example, Gynecology, Family/Internal Medicine, Radiology, Emergency Medicine). We also assessed EPL management type (medication, office procedure, OR procedure) by PEACE encounter category.

### Hypotheses

We hypothesized that PEACE utilization would be associated with a shorter care latency, and that patients who started care at PEACE would have the shortest care latency, followed by patients referred to PEACE as compared to controls, those patients who did not utilize PEACE. We hypothesized that a similar relationship would be observed between PEACE encounter group and our secondary outcome measures of care continuity and care efficiency. Finally, we sought to explore the relationship between patient demographic characteristics and PEACE encounter category.

### Statistical Analysis

We conducted a descriptive analysis of demographic variables by PEACE encounter category, and used multivariate logistic regression to compute odds ratios for PEACE utilization by patient demographic characteristics. We used the Chi-squared test of independence and one-way ANOVA or Kruskal-Wallis test where appropriate, to assess the relationship between PEACE utilization category and measures of care latency, efficiency, and continuity, with 2-tailed *p* < 0.05 as the cutoff for statistical significance. We also performed a survival analysis with a cox proportional hazard model of care latency by PEACE encounter category. All statistical analyses were conducted in Stata v18.0 (StataCorp, College Station, TX).

## Results

A total of 2,151 patients met inclusion criteria (Table 1), of whom 885 (41.1%) utilized PEACE during their EPL care. Among PEACE users, 246 (27.8%) initiated care at PEACE and 639 (72.2%) were referred after initial evaluation elsewhere. The cohort was racially and socioeconomically diverse, with 36.5% identifying as Black and 30.3% publicly insured.

**Table 1.** Cohort Demographics.

|  | **Overall** | **No PEACE** | **PEACE referral** | **PEACE start** | ***p*** |
| --- | --- | --- | --- | --- | --- |
|  | (n = 2,151) | (n = 1,266) | (n = 639) | (n = 246) |  |
| **Age, M(SD)** | 32.3 (5.5) | 32.3 (5.3) | 31.9 (5.6) | 32.8 (6.0) | 0.062* |
| **BMI, M(SD)** | 27.9 (7.1) | 27.9 (7.3) | 28.1 (6.8) | 27.4 (7.1) | 0.497* |
| **Race** |  |  |  |  | < 0.001** |
| White | 953 (44.3) | 584 (46.1) | 265 (41.5) | 104 (42.3) |  |
| Black | 785 (36.5) | 410 (32.4) | 272 (42.6) | 103 (41.9) |  |
| Mixed/Other | 279 (13) | 178 (14.1) | 71 (11.1) | 30 (12.2) |  |
| Missing | 134 (6.2) | 94 (7.4) | 31 (4.9) | 9 (3.7) |  |
| **Insurance** |  |  |  |  | < 0.001** |
| Private | 1397 (64.9) | 866 (68.4) | 370 (57.9) | 161 (65.5) |  |
| Public | 652 (30.3) | 335 (26.5) | 236 (36.9) | 81 (32.9) |  |
| Uninsured/Unknown | 102 (4.7) | 65 (5.1) | 33 (5.2) | 4 (1.6) |  |

PEACE, Pregnancy Early Access Center; EPL, early pregnancy loss; BMI, Body Mass Index (kg/m<sup>2</sup>)
Data are n (%).
\*ANOVA.
\*\*The Fisher exact test.

Care latency (Table 2) differed significantly by PEACE utilization category (p<0.001). Patients who initiated care at PEACE had the shortest median care latency at 0 days (IQR 0–2), compared with 5 days (IQR 0–13) among patients who never used PEACE and 8 days (IQR 4– 15) among patients referred to PEACE. In adjusted Cox regression, patients who initiated care at PEACE began treatment more than twice as quickly as patients who never used PEACE (aHR 2.36; 95% CI, 2.05–2.71), while patients referred to PEACE had longer time to treatment initiation than those who never used PEACE (aHR 0.83; 95% CI, 0.75–0.92). Care efficiency and continuity also differed by utilization category. Patients who initiated care at PEACE had fewer clinical encounters than patients referred to PEACE or those who never used PEACE: 2 encounters (IQR 1–3), 3 encounters (IQR 2–4), and 3 encounters (IQR 3–5), respectively (p<0.001). Patients who initiated care at PEACE also encountered fewer care teams: 4.5 teams (IQR 4–7), compared with 5 teams (IQR 5–8) among referred patients and 7 teams (IQR 5–9) among patients who never used PEACE (p<0.001).

**Table 2.**
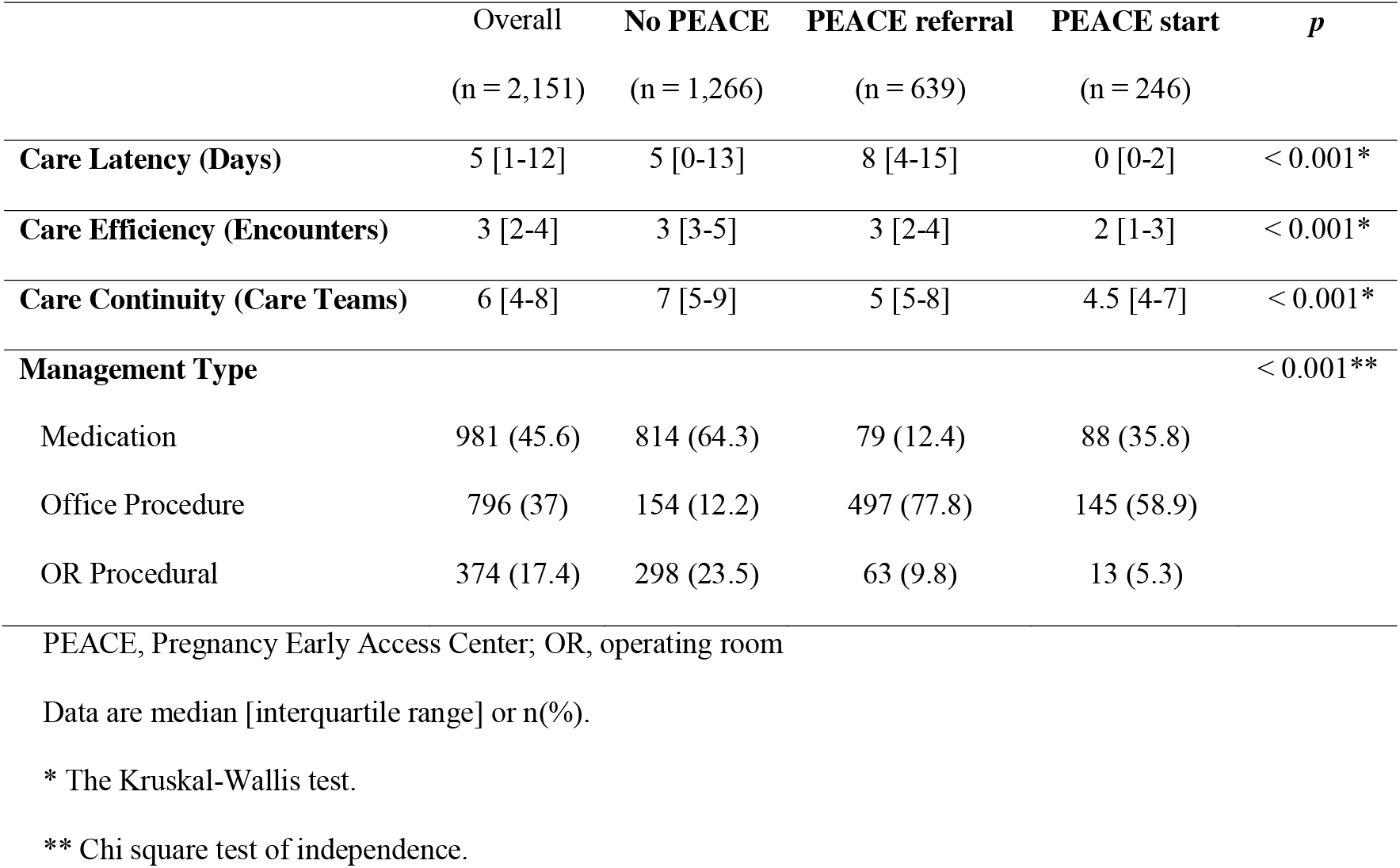
Comparison of outcomes by PEACE utilization category.

|  | Overall | No PEACE | PEACE referral | PEACE start | *p* |
| --- | --- | --- | --- | --- | --- |
|  | (n = 2,151) | (n = 1,266) | (n = 639) | (n = 246) |  |
| **Care Latency (Days)** | 5 [1-12] | 5 [0-13] | 8 [4-15] | 0 [0-2] | < 0.001* |
| **Care Efficiency (Encounters)** | 3 [2-4] | 3 [3-5] | 3 [2-4] | 2 [1-3] | < 0.001* |
| **Care Continuity (Care Teams)** | 6 [4-8] | 7 [5-9] | 5 [5-8] | 4.5 [4-7] | < 0.001* |
| **Management Type** |  |  |  |  | < 0.001** |
| Medication | 981 (45.6) | 814 (64.3) | 79 (12.4) | 88 (35.8) |  |
| Office Procedure | 796 (37) | 154 (12.2) | 497 (77.8) | 145 (58.9) |  |
| OR Procedural | 374 (17.4) | 298 (23.5) | 63 (9.8) | 13 (5.3) |  |

PEACE, Pregnancy Early Access Center; OR, operating room
Data are median [interquartile range] or n(%).
\* The Kruskal-Wallis test.
\*\* Chi square test of independence.

Among patients referred to PEACE (Table 3), time from initial symptom visit to PEACE encounter was strongly associated with care latency. Patients reaching PEACE within 7 days had a median care latency of 4 days (IQR 2–6), compared with 15 days (IQR 10–22) among those reaching PEACE after more than 7 days.

**Table 3.**
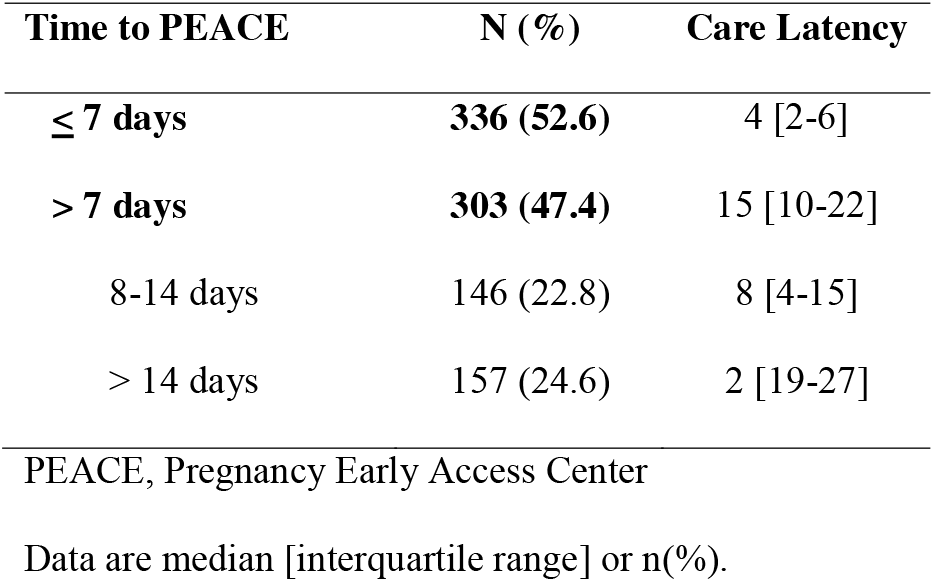
Comparison of care latency by the time between initial symptom visit and PEACE encounter for patients referred to PEACE.

| Time to PEACE | N (%) | Care Latency |
| --- | --- | --- |
| $\leq 7$ days | 336 (52.6) | 4 [2-6] |
| $> 7$ days | 303 (47.4) | 15 [10-22] |
| 8-14 days | 146 (22.8) | 8 [4-15] |
| $> 14$ days | 157 (24.6) | 2 [19-27] |

PEACE, Pregnancy Early Access Center
Data are median [interquartile range] or n(%).

Management type differed significantly across groups (p<0.001). Patients who started care at PEACE or were referred to PEACE were more likely to receive office-based uterine aspiration than patients who never used PEACE. Patients who never used PEACE were more likely to receive medication management or OR-based procedural management.

## Discussion

In this retrospective cohort study of patients undergoing active management of EPL, those who initiated care with the Pregnancy Early Access Center (PEACE) experienced shorter time to treatment initiation, fewer clinical encounters, and greater continuity of care compared with patients who did not access PEACE. These findings suggest that direct access to specialized ambulatory early pregnancy care may streamline EPL management and reduce fragmentation of care delivery,

The most notable finding was the marked reduction in care latency among patients who initiated care at PEACE, with a median latency of 0 days compared with 5 days among patients who never utilized PEACE. This association remained significant after adjustment for demographic factors. Patients who initiated care at PEACE also encountered fewer specialty teams and required fewer clinical visits, suggesting that PEACE may reduce both logistical barriers and care fragmentation during a highly vulnerable period. These findings are consistent with prior literature demonstrating improved efficiency, patient satisfaction, and quality of care in early pregnancy assessment units and specialized ambulatory pregnancy clinics [14–17].

In contrast, patients referred to PEACE after initial evaluation elsewhere experienced the longest care latency. This likely reflects delays associated with referral workflows, scheduling barriers, and transitions between care settings rather than inefficiency within PEACE itself. Supporting this interpretation, referred patients who reached PEACE within 7 days experienced substantially shorter care latency than those with longer referral intervals. Together, these findings suggest that the benefits of specialized early pregnancy care may depend not only on clinic availability, but also on timely and direct access pathways.

Our findings should be considered in the context of clinical decision-making and patient preference. Because EPL management is preference-sensitive [22]., patients may appropriately undergo a period of diagnostic uncertainty or expectant management before choosing active treatment [1]. In ur health system, PEACE serves as a referral center for office-based uterine aspiration, which may explain the high proportion of referred patients undergoing procedural management. Importantly, however, patients who initiated care at PEACE were also substantially more likely to receive office-based management than patients who never utilized PEACE, suggesting that direct access to specialized ambulatory care may expand access to preferred management options outside of the emergency department or operating room.

These findings are particularly relevant within the evolving reproductive healthcare landscape in the United States. Early pregnancy complications frequently arise before patients establish prenatal care, and many individuals rely on emergency departments for evaluation and management. Although ED presentation for EPL symptoms is common[23], emergency department-based EPL care is often associated with fragmented follow-up and lower patient satisfaction[4,23]. n the post-Dobbs environment, increasing fragmentation of reproductive healthcare access may further exacerbate delays in timely pregnancy loss care, particularly for historically underserved populations. Specialized ambulatory early pregnancy access models such as PEACE may represent an important strategy to improve timely, patient-centered care while reducing reliance on emergency departments, especially for traditionally underserved patient populations [20].

The strengths of this study include its large sample size, diverse patient population, nclusion of multiple care settings within a large urban health system, and application of a novel quantitative means of assessing patient care experiences. everal limitations should be considered. First, this was a retrospective observational study within a single healthcare system, limiting causal inference and generalizability. Patients who accessed PEACE may differ systematically from those who did not, including in care preferences, symptom severity, or healthcare-seeking behavior. Second, because exact timing of EPL diagnosis could not be reliably determined from administrative data, we used initial presentation for concerning pregnancy symptoms as the beginning of the care interval. Although this approach may overestimate latency for some patients undergoing diagnostic evaluation or expectant management, this limitation likely affected all groups similarly. Finally, we were unable to capture care received outside of our healthcare system.

In conclusion, direct access to ambulatory early pregnancy care was associated with shorter time to EPL management, fewer clinical encounters, and greater continuity of care. Given increased workforce pressures and access concerns in the U.S., these findings suggest the broader implementation of the PEACE model may help improve equitable access to timely, patient-centered pregnancy loss care.

## Data Availability

The data generated and analyzed during the current study are not publicly available and will not be shared externally. Relevant findings are contained within the manuscript.

## Appendix 1.

| EPL Diagnosis Codes | Abnormal Pregnancy Symptom<br>Diagnosis Codes | Excluded Diagnosis Codes | Treatment Codes |
| --- | --- | --- | --- |
| O02.1 – missed abortion | O20.0 – threatened abortion | Z33.2 – induced abortion | 59820 – surgical treatment of missed abortion |
| O03.x – spontaneous abortion | O36.8x – pregnancy of uncertain viability | O04.x – complicated induced abortion | 59812 – surgical treatment of incomplete abortion |
|  | O20.9 – bleeding in early pregnancy | O07.x – failed attempted abortion |  |
|  | O26.8x – spotting in early pregnancy | O01.x – molar pregnancy |  |
|  |  | O00.x – ectopic pregnancy |  |
|  |  | O26.2x – recurrent pregnancy loss |  |
|  |  | O30.x – multiple gestation |  |

